# Results of a Community-Based Coronary CT Angiography Screening in Asymptomatic Adults With Cardiovascular Risk Factors: The Broward County Heart Project

**DOI:** 10.64898/2026.10.12.26365337

**Authors:** Antonio Lewis, F. Scott Ross, Jerry D. Estep, David A. Baran

## Abstract

**Background:** Coronary CT angiography (CCTA) and calcium scoring detect subclinical coronary atherosclerosis, but the results of community-based screening in asymptomatic adults with cardiovascular risk factors are poorly described.

**Objective:** To describe the results of the first phase of a county-funded CT screening program and their variation by demographics, health system, time, and ZIP code.

**Methods:** Retrospective analysis of aggregate reports from the Broward County Florida Heart Project (May 2023–September 2025). Eligible residents were those with insurance coverage, age 45–70 years (limited exceptions), free of known cardiovascular disease and symptoms, with at least one risk factor. Examinations at six hospitals in four health systems were classified by the program team as normal (Category 1), abnormal (Category 2), or very abnormal (Category 3), with a separate flag for other non-coronary abnormality. We used Wilson confidence intervals and chi-square, trend, and Mantel-Haenszel tests.

**Results:** Among 7,135 participants (58.3% men; 79.0% White, 8.0% Black, 17.2% Hispanic), 43.2% had Category 1, 45.4% Category 2, and 11.4% Category 3 results; 56.8% (95% CI, 55.7%–58.0%) had abnormal or very abnormal results. The proportion rose from 39.3% at 45–55 years to 78.5% at 71 or older (trend P<0.001) and was higher in men than women (68.9% vs 39.8%; odds ratio [OR], 3.36; 95% CI, 3.05–3.71). Rates were lower in Black than White participants (38.9% vs 58.6%; hospital-adjusted OR, 0.45; 95% CI, 0.38–0.54) and in Hispanic than non-Hispanic participants (51.0% vs 57.8%). Abnormal results ranged from 39.5% to 60.3% across health systems (P<0.001). Variation across the 50 highest-volume ZIP codes did not exceed chance (P=0.53).

**Conclusions:** More than half of insured, asymptomatic adults with risk factors screened in this community program had abnormal results, with steep age and sex gradients and unexplained variation across health systems.

## INTRODUCTION

Atherosclerotic cardiovascular disease remains a leading cause of death in the United States, and primary prevention relies largely on estimating risk from traditional risk factors. Such estimates cannot show directly whether a person already has coronary atherosclerosis. Coronary artery calcium (CAC) scoring is recommended in current guidelines to refine risk assessment in selected adults when treatment decisions are uncertain,^1^ and coronary CT angiography (CCTA) adds direct visualization of noncalcified plaque and luminal stenosis. Much of the trial evidence for CCTA, including SCOT-HEART and PROMISE, derived from symptomatic patients evaluated for suspected coronary artery disease.^2,3^

The utility and diagnostic yield of CCTA in a community cohort of asymptomatic patients is not well characterized. In the Swedish SCAPIS study, which used CCTA in 25,182 randomly invited adults aged 50 to 64 years without known coronary disease, 42.1% had any coronary atherosclerosis and 5.2% had stenosis of at least 50%.^4^ The Danish DANCAVAS trial randomized men aged 65 to 74 years to population screening that included CAC scoring and did not show a statistically significant reduction in all-cause mortality after a median of 5.6 years (hazard ratio, 0.95; 95% CI, 0.90–1.00).^5^ Less is known about the results of community-based CT screening offered to a diverse US population on the basis of individual risk factors in asymptomatic participants. Since coronary atherosclerosis is often silent until a first cardiac event, many adults at risk are unaware of their disease. Publicly supported community programs that offer screening outside the usual care setting may reach people who would not otherwise seek cardiovascular evaluation, raise community awareness of atherosclerotic cardiovascular disease (ASCVD), and help orient local health care toward prevention rather than treatment of advanced disease.

The Broward County Florida Heart Project was a county-funded program that offered CCTA or calcium scoring at no cost to county residents aged 45 to 70 years (with limited age exceptions) who had risk factors for coronary disease but no known cardiovascular disease or symptoms. Its first phase, run by a third-party administrator (SydCura) from May 2023 through September 2025, delivered examinations at six hospitals in four health systems. At the program’s conclusion the administrator produced program-level reports on participation and results. We used these reports to describe the results of Phase 1 and their variation by age, sex, race, ethnicity, health system, calendar period, and ZIP code of residence.

## METHODS

### Design and data sources

This was a retrospective cross-sectional analysis of aggregate program data, reported according to the STROBE guideline.^6^ The sources were two documents prepared by the administrator for Broward County: a final report covering May 1, 2023, through September 30, 2025 (dated January 23, 2026),^7^ and a monthly reporting deck with data collected through November 30, 2025.^8^ Neither contained individual-level data. On this basis, this work was exempt from Institutional Review Board approval.

### Program and participants

Residents applied through a county program website and completed a questionnaire. The administrator verified Broward County Florida residency and health insurance, collected informed consent and waiver forms approved by the County, screened applicants against criteria set by a Medical Advisory Committee, and scheduled eligible applicants at participating health systems. Applicants were required to be Broward County residents aged 45 to 70 years with active health insurance and at least one of the following: current smoking, diabetes, hypertension, high cholesterol, or a family history of coronary disease. Applicants were excluded if they reported a prior cardiac CT or calcium scoring, chest pain or shortness of breath, inhaler use, prior coronary stent placement, prior cardiac surgery, iodine contrast allergy, chemotherapy or immunotherapy, or a solid organ transplant. The program reported that 4.3% of participants had an age exception and 0.02% a medical exception.

Participants underwent CCTA (CPT 75574) or calcium scoring alone (CPT 75571) at one of six hospitals belonging to four health systems. To deidentify the participating institutions, we refer to the health systems as Sites A to D in descending order of participant volume. Results by individual hospital are not presented.

### Outcome classification

The program team classified each result as Category 1 (normal), Category 2 (abnormal), or Category 3 (very abnormal). A fourth flag, Category 4 (other abnormality, non-coronary), was recorded in addition to one of the first three categories. The program treated Categories 2 and 3 as requiring follow-up. The operational definitions of the categories, including any calcium score or stenosis thresholds, and the process by which examinations were assigned to them were not included in the reports, and the categories are not expressed in an accepted reporting system such as CAD-RADS 2.0.^9^ We therefore use the program’s labels throughout. The primary outcome of interest was an abnormal or very abnormal result (Category 2 or 3 versus Category 1). Secondary outcomes were a very abnormal result (Category 3 versus Categories 1 and 2) and the Category 4 flag.

### Participant characteristics and comparison groups

The reports tabulated results by age group (45–55, 56–65, 66–70, and 71 years or older), sex, race, ethnicity, health system, and service period (May 2023–April 2024, May 2024–April 2025, and May–September 2025). The final report also listed the 50 ZIP codes with the highest participation, with the number of participants in each and the number with Category 2 or 3 results.

### Statistical analysis

We calculated proportions with Wilson 95% confidence intervals. Proportions were compared across groups with Pearson chi-square tests, and ordered groups (age and service period) with the Cochran-Armitage test for trend. Odds ratios and Woolf 95% confidence intervals were calculated for sex and ethnicity. For race, the report also provided results by hospital, which allowed us to estimate a Mantel-Haenszel odds ratio for Black versus White participants stratified by the six hospitals and to test for heterogeneity across hospitals. Variation across ZIP codes was assessed with a chi-square test of homogeneity. Tests were two-sided with a significance level of 0.05, without adjustment for multiple comparisons, and secondary and subgroup analyses should be considered exploratory. Because only aggregate data were available, we could not adjust for age, sex, or risk factors jointly. Where the same quantity appeared in more than one table with small differences (no more than four participants in any cell), we used the table that matched the analysis. Analyses used Python 3 (SciPy 1.13.1 and statsmodels 0.14.6).

## RESULTS

### Program participation

From May 2023 through September 2025 the program website recorded 24,698 visits and 10,358 completed applications, and 5,343 applicants did not meet eligibility criteria (Table 1). A total of 7,664 examinations were scheduled, 7,131 were completed and paid, and 7,135 participants had results. The overall no-show rate was 3.8%. Physicians and other medical professionals (5,311 responses) and family members or friends (4,941) were the most commonly reported referral sources. The most frequent reasons for ineligibility, which were not mutually exclusive, were chest pain or shortness of breath (1,466; 27.4%); age outside 45 to 70 years (1,153; 21.6%); a prior cardiac CT or calcium score (1,075; 20.1%); no active health insurance (710; 13.3%); and inhaler use (583; 10.9%).

**Table 1.**
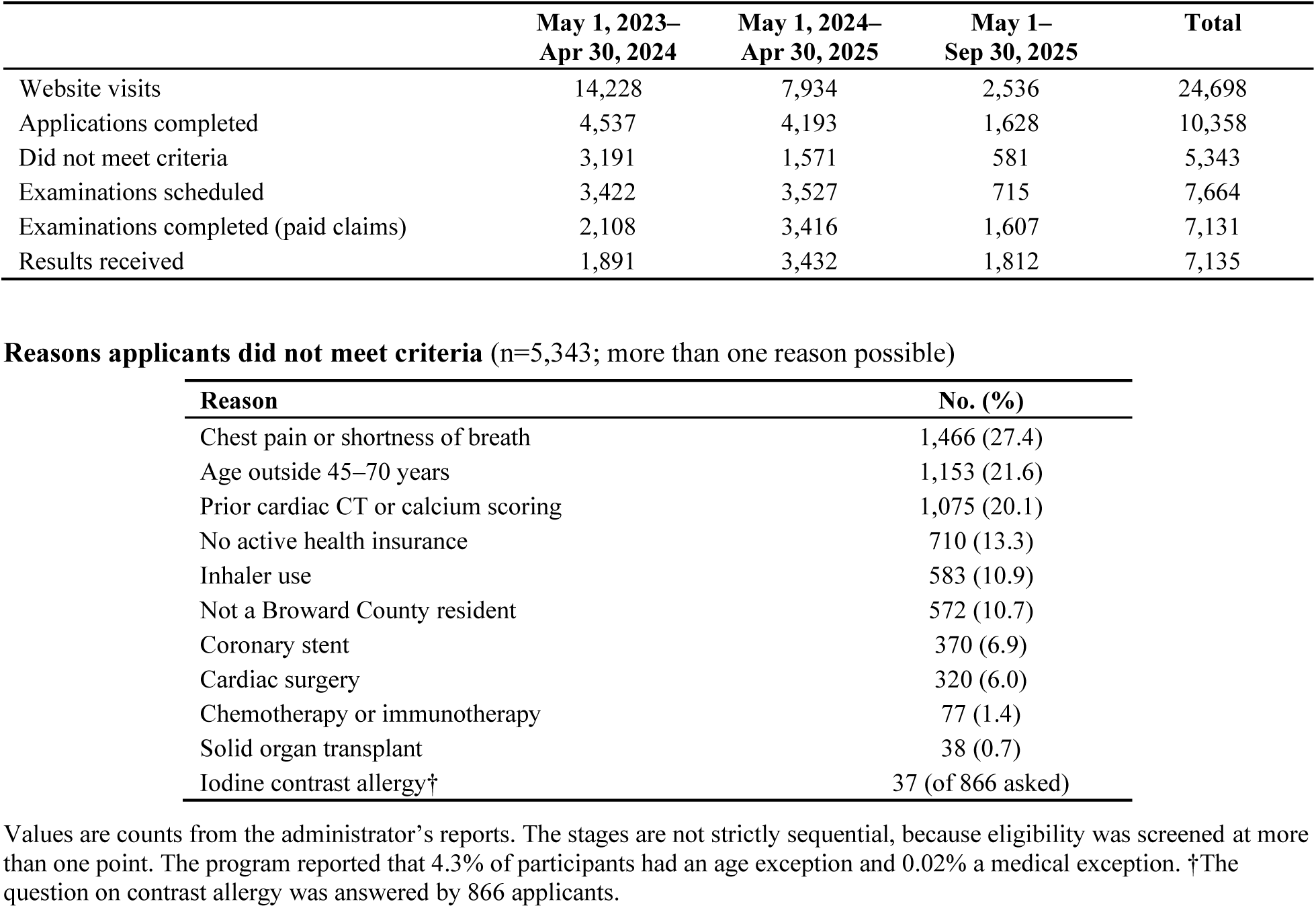
Program participation and reasons applicants did not meet eligibility criteria, May 2023–September 2025.

|  | May 1, 2023–<br>Apr 30, 2024 | May 1, 2024–<br>Apr 30, 2025 | May 1–<br>Sep 30, 2025 | Total |
| --- | --- | --- | --- | --- |
| Website visits | 14,228 | 7,934 | 2,536 | 24,698 |
| Applications completed | 4,537 | 4,193 | 1,628 | 10,358 |
| Did not meet criteria | 3,191 | 1,571 | 581 | 5,343 |
| Examinations scheduled | 3,422 | 3,527 | 715 | 7,664 |
| Examinations completed (paid claims) | 2,108 | 3,416 | 1,607 | 7,131 |
| Results received | 1,891 | 3,432 | 1,812 | 7,135 |

**Reasons applicants did not meet criteria** (n=5,343; more than one reason possible)
| Reason | No. (%) |
| --- | --- |
| Chest pain or shortness of breath | 1,466 (27.4) |
| Age outside 45–70 years | 1,153 (21.6) |
| Prior cardiac CT or calcium scoring | 1,075 (20.1) |
| No active health insurance | 710 (13.3) |
| Inhaler use | 583 (10.9) |
| Not a Broward County resident | 572 (10.7) |
| Coronary stent | 370 (6.9) |
| Cardiac surgery | 320 (6.0) |
| Chemotherapy or immunotherapy | 77 (1.4) |
| Solid organ transplant | 38 (0.7) |
| Iodine contrast allergy† | 37 (of 866 asked) |

Values are counts from the administrator’s reports. The stages are not strictly sequential, because eligibility was screened at more than one point. The program reported that 4.3% of participants had an age exception and 0.02% a medical exception. †The question on contrast allergy was answered by 866 applicants.

### Participant characteristics

Of the 7,135 participants with results, 2,084 (29.2%) were aged 45 to 55 years, 3,527 (49.4%) were 56 to 65, 1,205 (16.9%) were 66 to 70, and 317 (4.4%) were 71 or older; age was under 45 or missing for 2. Men accounted for 4,163 (58.3%) and women for 2,966 (41.6%); sex was unknown for 6. Participants were 79.0% White, 8.0% Black or African American, 2.7% Asian, 5.3% some other race, and 4.9% unknown race; 17.2% were Hispanic or Latino. Site A contributed 4,010 participants (56.2%), Site B 1,869 (26.2%), Site C 691 (9.7%), and Site D 565 (7.9%) (Table 2).

**Table 2.** Participant characteristics and program result categories (N=7,135)

| Characteristic | No. (%) of participants | Category 1 normal, No. (%) | Category 2 abnormal, No. (%) | Category 3 very abnormal, No. (%) | Category 2 or 3, % (95% CI) | P value* |
| --- | --- | --- | --- | --- | --- | --- |
| **All participants** | **7,135 (100)** | **3,081 (43.2)** | **3,241 (45.4)** | **813 (11.4)** | **56.8 (55.7–58.0)** |  |
| **Age group, y** |  |  |  |  |  | **<0.001†** |
| 45–55 | 2,084 (29.2) | 1,264 (60.7) | 718 (34.5) | 102 (4.9) | 39.3 (37.3–41.5) |  |
| 56–65 | 3,527 (49.4) | 1,430 (40.5) | 1,660 (47.1) | 437 (12.4) | 59.5 (57.8–61.1) |  |
| 66–70 | 1,205 (16.9) | 318 (26.4) | 678 (56.3) | 209 (17.3) | 73.6 (71.0–76.0) |  |
| 71+ | 317 (4.4) | 68 (21.5) | 184 (58.0) | 65 (20.5) | 78.5 (73.7–82.7) |  |
| **Sex** |  |  |  |  |  | **<0.001** |
| Male | 4,163 (58.3) | 1,294 (31.1) | 2,224 (53.4) | 645 (15.5) | 68.9 (67.5–70.3) |  |
| Female | 2,966 (41.6) | 1,787 (60.2) | 1,012 (34.1) | 167 (5.6) | 39.8 (38.0–41.5) |  |
| **Race** |  |  |  |  |  | **<0.001** |
| White | 5,637 (79.0) | 2,335 (41.4) | 2,629 (46.6) | 673 (11.9) | 58.6 (57.3–59.9) |  |
| Black or African American | 571 (8.0) | 349 (61.1) | 194 (34.0) | 28 (4.9) | 38.9 (35.0–42.9) |  |
| Asian | 195 (2.7) | 92 (47.2) | 82 (42.1) | 21 (10.8) | 52.8 (45.8–59.7) |  |
| Some other race | 381 (5.3) | 176 (46.2) | 157 (41.2) | 48 (12.6) | 53.8 (48.8–58.7) |  |
| Unknown | 351 (4.9) | 129 (36.8) | 179 (51.0) | 43 (12.3) | 63.2 (58.1–68.1) |  |
| **Ethnicity** |  |  |  |  |  | **<0.001‡** |
| Hispanic or Latino | 1,226 (17.2) | 601 (49.0) | 520 (42.4) | 105 (8.6) | 51.0 (48.2–53.8) |  |
| Not Hispanic or Latino | 5,827 (81.7) | 2,458 (42.2) | 2,672 (45.9) | 697 (12.0) | 57.8 (56.5–59.1) |  |
| Unknown | 82 (1.1) | 22 (26.8) | 49 (59.8) | 11 (13.4) | 73.2 (62.7–81.6) |  |
| **Health system** |  |  |  |  |  | **<0.001** |
| Site A | 4,010 (56.2) | 1,592 (39.7) | 1,842 (45.9) | 576 (14.4) | 60.3 (58.8–61.8) |  |
| Site B | 1,869 (26.2) | 769 (41.1) | 972 (52.0) | 128 (6.8) | 58.9 (56.6–61.1) |  |
| Site C | 691 (9.7) | 418 (60.5) | 209 (30.2) | 64 (9.3) | 39.5 (35.9–43.2) |  |
| Site D | 565 (7.9) | 301 (53.3) | 219 (38.8) | 45 (8.0) | 46.7 (42.6–50.8) |  |
| **Service period** |  |  |  |  |  | **0.014†** |
| May 2023–Apr 2024 | 1,891 (26.5) | 783 (41.4) | 954 (50.4) | 154 (8.1) | 58.6 (56.4–60.8) |  |
| May 2024–Apr 2025 | 3,432 (48.1) | 1,475 (43.0) | 1,502 (43.8) | 455 (13.3) | 57.0 (55.4–58.7) |  |
| May–Sep 2025 | 1,812 (25.4) | 823 (45.4) | 785 (43.3) | 204 (11.3) | 54.6 (52.3–56.9) |  |

\*Chi-square test of the proportion with Category 2 or 3 versus Category 1 across the groups shown, unless otherwise indicated.
†Cochran-Armitage test for trend. ‡Hispanic versus non-Hispanic participants. Age excludes 2 participants younger than 45 years or with missing age; sex excludes 6 participants with unknown sex (5 Category 2, 1 Category 3). Percentages in the category columns are row percentages. Health systems are coded A to D in descending order of participant volume. Race and ethnicity were recorded as reported to the program.

### Overall results

Overall, 3,081 participants (43.2%) had Category 1 results, 3,241 (45.4%) had Category 2, and 813 (11.4%) had Category 3 (Figure 1). An abnormal or very abnormal result (Category 2 or 3) was therefore present in 4,054 participants (56.8%; 95% CI, 55.7%–58.0%). Table 4 and the Central Illustration summarize the principal findings.

**Figure 1.**
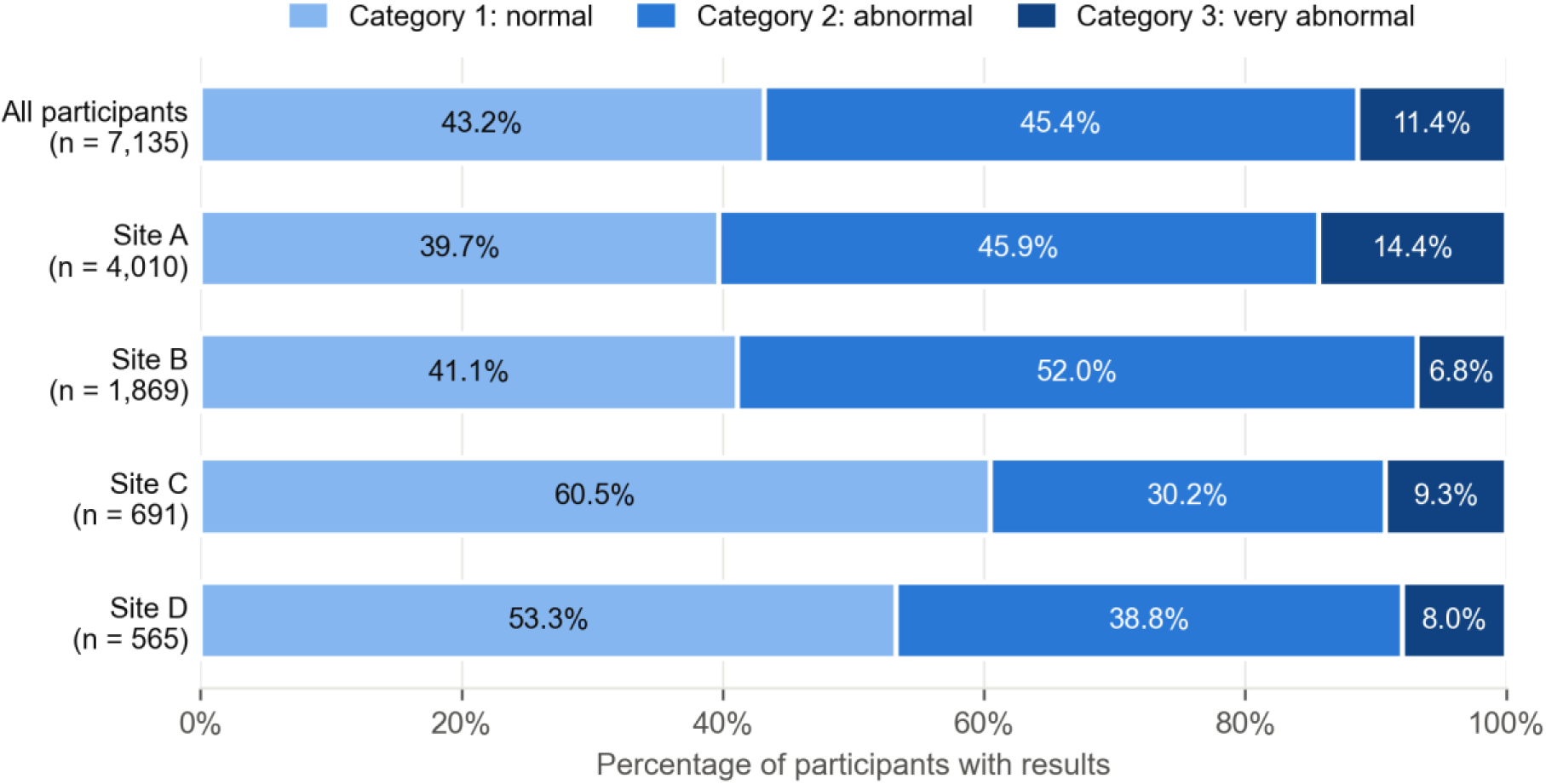
Distribution of program result categories among 7,135 participants, overall and by health system. Categories were assigned by the program: Category 1, normal; Category 2, abnormal; Category 3, very abnormal. Sites A to D are health systems in descending order of participant volume. Percentages may not sum to 100 because of rounding.

### Age and sex

The proportion with an abnormal or very abnormal result increased with age, from 39.3% (95% CI, 37.3%–41.5%) at 45 to 55 years to 59.5% at 56 to 65, 73.6% at 66 to 70, and 78.5% (95% CI, 73.7%–82.7%) at 71 years or older (trend P<0.001; Table 2, Figure 2). The proportion with a very abnormal result rose from 4.9% to 12.4%, 17.3%, and 20.5% (trend P<0.001). Men were more likely than women to have an abnormal or very abnormal result (68.9% vs 39.8%; OR, 3.36; 95% CI, 3.05–3.71) and a very abnormal result (15.5% vs 5.6%; OR, 3.07; 95% CI, 2.57–3.67; both P<0.001).

**Figure 2.**
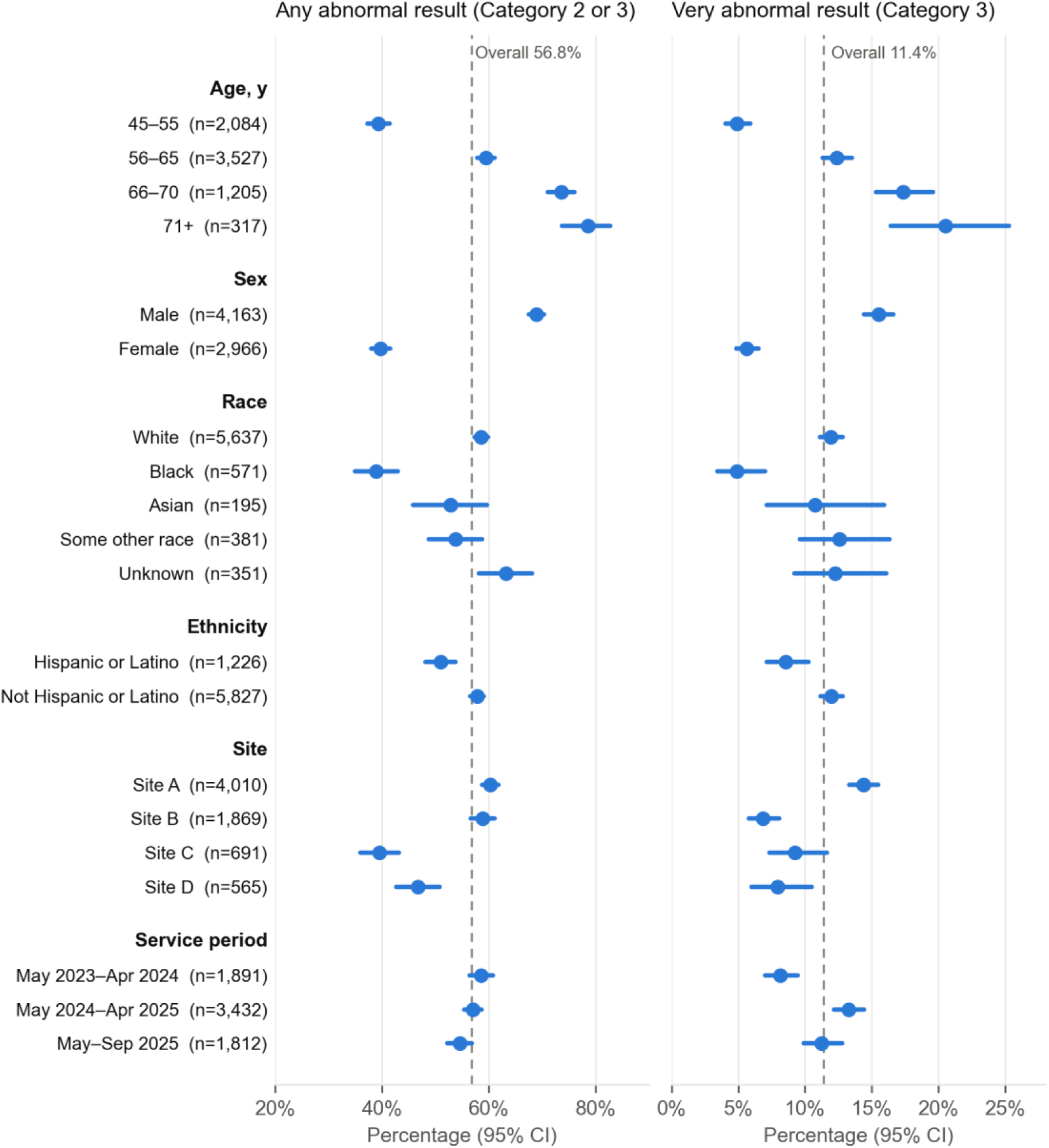
Proportion of participants with an abnormal or very abnormal result (Category 2 or 3; left) and a very abnormal result (Category 3; right) by participant characteristic, health system, and service period. Points are proportions and horizontal lines are Wilson 95% confidence intervals; dashed vertical lines mark the overall proportion. Age excludes 2 participants younger than 45 years or with missing age, sex excludes 6 with unknown sex, and ethnicity excludes 82 with unknown ethnicity.

### Race and ethnicity

Black participants had lower rates of abnormal or very abnormal results than White participants (38.9% [95% CI, 35.0%–42.9%] vs 58.6% [95% CI, 57.3%–59.9%]; crude OR, 0.45; 95% CI, 0.38–0.54). The association was unchanged after stratification by hospital (Mantel-Haenszel OR, 0.45; 95% CI, 0.38–0.54; P<0.001), with no evidence of heterogeneity across hospitals (P=0.50). After the same stratification, the odds among Asian participants did not differ significantly from those among White participants (OR, 0.81; 95% CI, 0.60–1.10). Hispanic participants had lower rates than non-Hispanic participants of abnormal or very abnormal results (51.0% vs 57.8%; OR, 0.76; 95% CI, 0.67–0.86; P<0.001) and of very abnormal results (8.6% vs 12.0%).

### Health system and calendar period

Abnormal or very abnormal results varied across the four health systems, from 39.5% at Site C to 60.3% at Site A (χ²=130.8, 3 df; P<0.001), and very abnormal results ranged from 6.8% at Site B to 14.4% at Site A (χ²=83.0, 3 df; P<0.001; Table 2, Figure 2). Site B had a high proportion of Category 2 results (52.0%) but the lowest proportion of Category 3. Across the three service periods, the proportion with an abnormal or very abnormal result declined modestly (58.6%, 57.0%, and 54.6%; trend P=0.01), whereas the proportion with a very abnormal result rose from 8.1% to 13.3% and was 11.3% in the final period (P<0.001).

### Other non-coronary abnormality

Category 4 (other non-coronary abnormality) was identified in 362 participants (5.1%; 95% CI, 4.6%–5.6%), including 246 of those with Category 1 results, 100 with Category 2, and 16 with Category 3 (Table 3). Its frequency varied by health system, from 2.7% at Site D to 7.8% at Site C (P<0.001), and rose from 3.8% in the first service period to 5.5% and 5.5% in the later periods (P=0.01). It was identified more often among Black participants (8.2%) than White participants (4.8%; P=0.004 across race groups).

**Table 3.** Category 4 (other abnormality, non-coronary) flag.

| **Group** | **Flagged / total** | **% (95% CI)** | **P value\*** |
| --- | --- | --- | --- |
| **All participants** | **362 / 7,135** | **5.1 (4.6–5.6)** |  |
| **Coexisting coronary category** |  |  |  |
| Category 1 (normal) | 246 / 3,081 | 8.0 (7.1–9.0) |  |
| Category 2 (abnormal) | 100 / 3,241 | 3.1 (2.5–3.7) |  |
| Category 3 (very abnormal) | 16 / 813 | 2.0 (1.2–3.2) |  |
| **Health system** |  |  | **<0.001** |
| Site A | 219 / 4,010 | 5.5 (4.8–6.2) |  |
| Site B | 74 / 1,869 | 4.0 (3.2–4.9) |  |
| Site C | 54 / 691 | 7.8 (6.0–10.1) |  |
| Site D | 15 / 565 | 2.7 (1.6–4.3) |  |
| **Service period** |  |  | **0.014** |
| May 2023–Apr 2024 | 72 / 1,891 | 3.8 (3.0–4.8) |  |
| May 2024–Apr 2025 | 190 / 3,432 | 5.5 (4.8–6.4) |  |
| May–Sep 2025 | 100 / 1,812 | 5.5 (4.6–6.7) |  |
| **Race†** |  |  | **0.004** |
| White | 252 / 5,217 | 4.8 (4.3–5.4) |  |
| Black | 44 / 537 | 8.2 (6.2–10.8) |  |
| Asian | 5 / 182 | 2.7 (1.2–6.3) |  |
| Other | 13 / 347 | 3.7 (2.2–6.3) |  |
| Unknown or refused | 48 / 851 | 5.6 (4.3–7.4) |  |

\*Chi-square test across the groups shown. †Race groups in this table come from the administrator's facility-by-race tabulations, which classify some participants as unknown or refused who are classified by race in the main race table (Table 2); denominators therefore differ slightly from Table 2. Category 4 was flagged in addition to Category 1, 2, or 3.

**Table 4.**
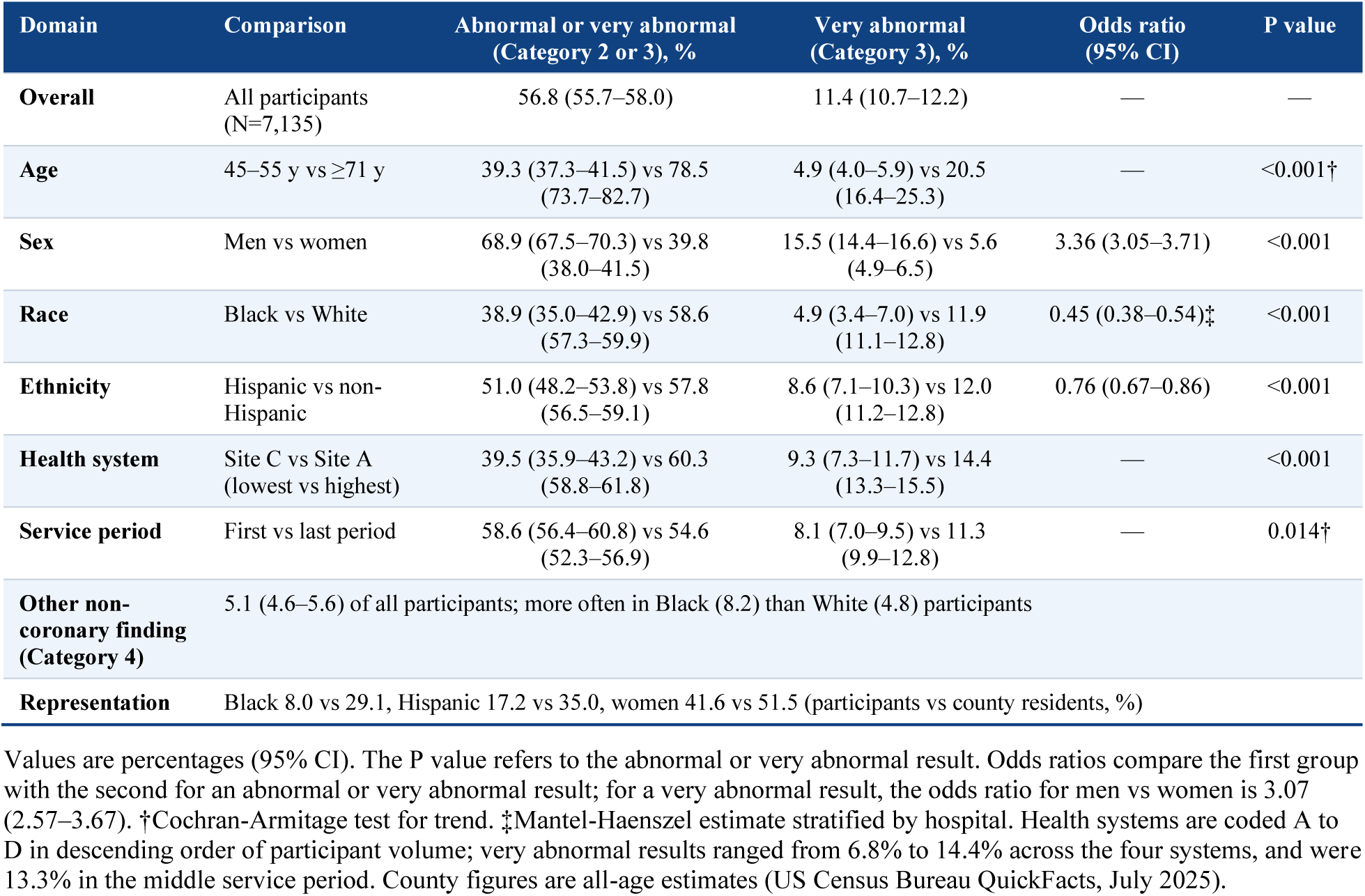
Summary of principal findings.

### ZIP code of residence

The 50 ZIP codes with the highest participation accounted for 6,796 participants (95.2% of 7,135). In these ZIP codes the proportion with an abnormal or very abnormal result ranged from 47.4% (ZIP code 33028; 45 of 95) to 69.8% (ZIP code 33009; 37 of 53), with a median of 57.3% (interquartile range, 54.1%–59.7%; Figure 3). This variation was no greater than expected by chance (χ²=47.6, 49 df; P=0.53). The ZIP code highlighted by the administrator for its number of abnormal results (33308; 220 of 361) had a proportion of 60.9%, similar to the pooled proportion of 57.1%.

**Figure 3.**
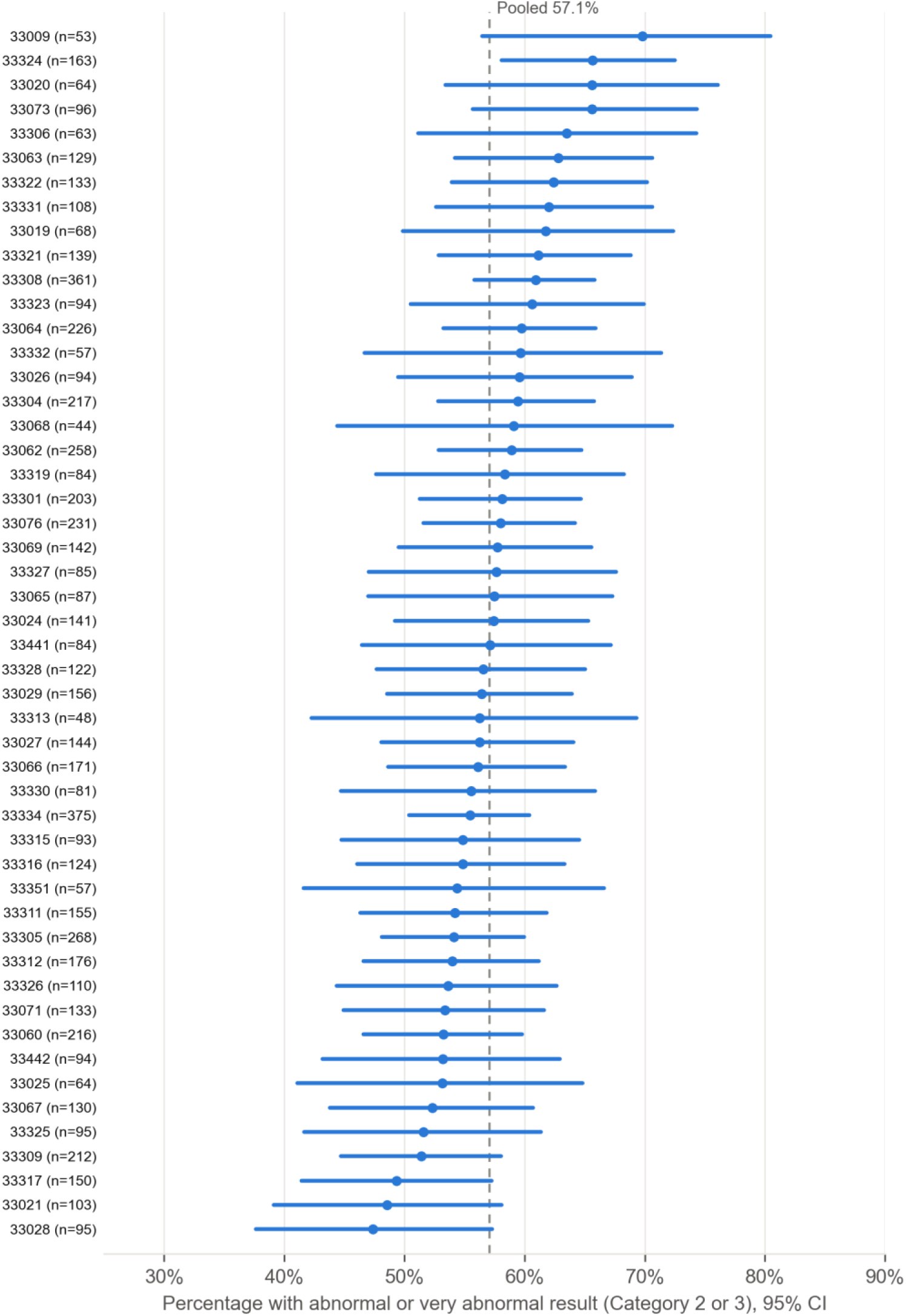
Proportion with an abnormal or very abnormal result (Category 2 or 3) in the 50 ZIP codes with the highest participation (6,796 participants), sorted by proportion. Points are proportions with Wilson 95% confidence intervals; the dashed line is the pooled proportion (57.1%). Variation across ZIP codes was no greater than expected by chance (χ²=47.6, 49 df; P=0.53).

## DISCUSSION

In this analysis of 7,135 asymptomatic, insured adults with at least one cardiovascular risk factor who were screened with CT in a county-funded program, 56.8% had an abnormal or very abnormal result and 11.4% had a very abnormal result. The proportion with abnormal results rose steeply with age and was much higher in men than in women. Results differed across four participating health systems, and lower rates were seen in Black and Hispanic participants. Among the 50 ZIP codes with the highest participation, we found no evidence that abnormal results were concentrated in particular areas.

These figures are hard to compare directly with other reports, because the program’s categories are not defined in terms of accepted CT reporting systems such as CAD-RADS 2.0^9^ and the population was selected on risk factors. For reference, SCAPIS found any atherosclerosis on CCTA in 42.1% of randomly invited adults aged 50 to 64 years,^4^ and the proportions with Category 2 or 3 results in our participants aged 45 to 55 years (39.3%) and 56 to 65 years (59.5%) are of broadly similar magnitude, although the populations, imaging protocols, and definitions differ. The age and sex gradients are consistent with the distribution of coronary calcium in the Multi-Ethnic Study of Atherosclerosis (MESA), in which calcium prevalence and amount were higher in men and increased steadily with age.^10^

Lower rates of abnormal results among Black and Hispanic participants also parallel MESA, in which the distribution of calcium differed by race and ethnicity, although the pattern varied by age and sex.^10^ Since calcium scores predicted coronary events in each of four racial and ethnic groups,^11^ a lower prevalence in a group does not imply that screening is less informative for its members. Our comparisons could not be adjusted for age, sex, or risk-factor profile, but stratifying by hospital did not change the difference between Black and White participants.

Delivering more than 7,000 examinations in 29 months across four health systems indicates that a county-led program can achieve substantial reach; whether that reach extends to the residents at highest risk is a separate question. Participation differed markedly from the county population. Countywide, 29.1% of residents are Black, 35.0% are Hispanic or Latino, and 51.5% are female,^12^ compared with 8.0%, 17.2%, and 41.6% of participants. The county figures include all ages, so the comparison is imperfect, but the gaps are large. The requirements for health insurance and for a risk factor, and reliance on referral through physicians and word of mouth, which were the most commonly reported referral sources in the administrator’s data, may have contributed. Targeted outreach to women and to Black and Hispanic residents may be needed for a program of this kind to serve the whole county.

Variation across health systems was large, especially for very abnormal results (6.8% to 14.4%), and is unlikely to be sampling error. Possible explanations include differences in the age, sex, and risk-factor profile of participants, referral patterns, and differences in how examinations were read or assigned to categories. These cannot be separated with aggregate data. The increase in very abnormal results between the first and second service years and the increase in the Category 4 flag may likewise reflect changes in the participant mix, the mix of sites, or classification practice. We regard these differences as hypotheses that require patient-level data for proper study.

The administrator suggested that future outreach could focus on ZIP codes with higher cardiovascular risk. Our analysis gives little support to ZIP code alone as a guide to the yield of screening: variation in abnormal results across the 50 highest-volume ZIP codes was compatible with chance. This finding is limited, because each ZIP code contributed only 44 to 375 participants, the comparison was unadjusted, and we had no data on area-level factors such as food access or deprivation. It does not exclude geographic variation that would appear in patient-level data.

The program treated Category 2 and 3 results as requiring follow-up, which applied to more than half of participants. This is a classification of results and not a record of care. We cannot say how many participants began preventive therapy, had further testing or revascularization, or benefited. Detection of subclinical disease does not by itself establish benefit, and a large randomized trial of population screening that included CAC scoring did not show a statistically significant mortality reduction.^5^ The proportions reported here indicate the scale of downstream evaluation and preventive treatment that a program of this design can generate.

Strengths of this analysis include the large number of participants, the use of common eligibility criteria across four health systems, and a community setting rather than an academic referral population. However, it has several limitations. First, only aggregate data were available, so we could not adjust for confounders or examine interactions, and the analyses of race, ethnicity, site, and ZIP code are unadjusted. Second, the result categories are program-defined, undefined in the reports, and not mapped to CAD-RADS or calcium scores; interpretation was performed at each site without central adjudication, and calcium-scoring-only examinations could not be separated from CCTA. Third, the reports did not describe the risk-factor profile of the scanned participants. The questionnaire data in the reports refer to applicants who did not qualify. Fourth, participants were self-selected, insured, and often physician-referred, and the source population is not defined, so the results cannot be generalized to the county or to uninsured adults. Fifth, race and ethnicity were missing or unknown for some participants, and the administrator’s tabulations differed slightly between tables. Sixth, follow-up care and outcomes were not measured.

Patient-level analysis of the program’s images, reports, and clinical records, using standardized definitions such as CAD-RADS and calcium scores and including risk-factor data, would allow adjustment for case mix and a direct assessment of variation across sites and areas, and such work is planned.

## CONCLUSIONS

In a community CT screening program for asymptomatic, insured adults aged 45 years and older with cardiovascular risk factors, more than half of participants had an abnormal result and about one in nine had a very abnormal result. The proportions rose steeply with age, were higher in men, were lower in Black and Hispanic participants, and varied substantially across health systems without a clear explanation. Black, Hispanic, and female residents were under-represented. By identifying previously unrecognized abnormal findings in adults with no known cardiovascular disease and no symptoms, county-supported screening of this kind may raise awareness of ASCVD and help shift the local health care system toward prevention; whether it improves outcomes remains to be seen.

## Conflict of interest

The authors have no conflicts of interest to disclose.

## Data Availability

All data produced in the present study are available upon reasonable request to the authors

---

**Central Illustration.**
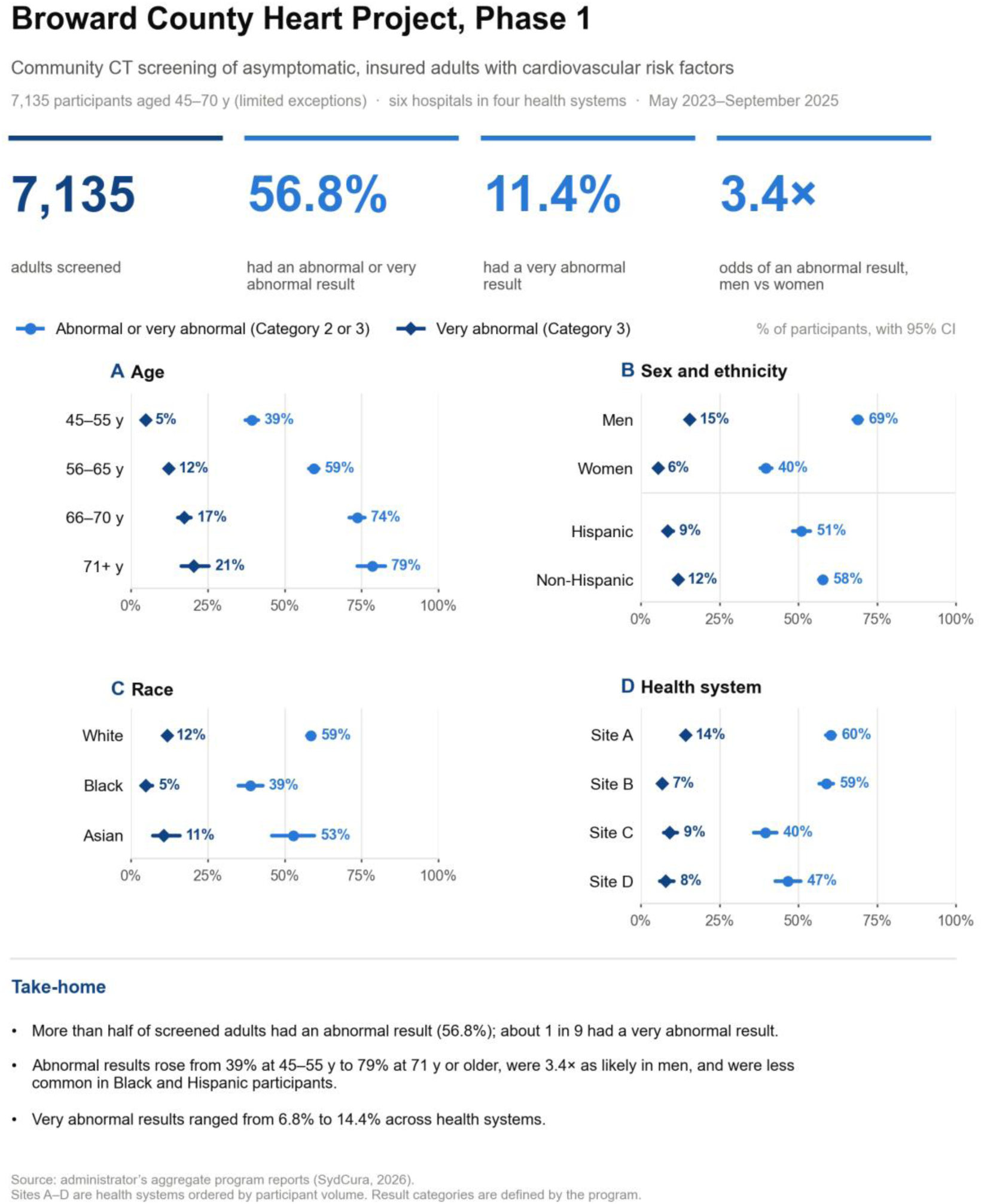
Principal findings of Phase 1 of the Broward County Heart Project (N=7,135). Panels A to D show the percentage of participants with an abnormal or very abnormal result (Category 2 or 3; circles) and a very abnormal result (Category 3; diamonds), with Wilson 95% confidence intervals, by age, by sex and ethnicity, by race, and by health system (Sites A to D, in descending order of volume).

## Notes

### Competing Interest Statement

The authors have declared no competing interest.

### Author Declarations

This manuscript reports aggregate, program-level data from the Broward County Heart Project, provided by SydCura Health Solutions, the Countys program administrator. The authors did not access patient-level data, identifiable information, or medical records for this analysis, and no ethics committee or institutional review board review was obtained for this manuscript. A separate protocol for patient-level analysis of the same program is under review by the Cleveland Clinic Institutional Review Board and is not part of this work. No data from that protocol are reported here.

### Funding

- Broward County Government

## REFERENCES

1. Arnett DK, Blumenthal RS, Albert MA, et al. 2019 ACC/AHA guideline on the primary prevention of cardiovascular disease: a report of the American College of Cardiology/American Heart Association Task Force on Clinical Practice Guidelines. Circulation. 2019;140(11):e596–e646. doi:10.1161/CIR.0000000000000678

2. SCOT-HEART Investigators. CT coronary angiography in patients with suspected angina due to coronary heart disease (SCOT-HEART): an open-label, parallel-group, multicentre trial. Lancet. 2015;385(9985):2383–2391. doi:10.1016/S0140-6736(15)60291-4

3. Douglas PS, Hoffmann U, Patel MR, et al. Outcomes of anatomical versus functional testing for coronary artery disease. N Engl J Med. 2015;372(14):1291–1300. doi:10.1056/NEJMoa1415516

4. Bergström G, Persson M, Adiels M, et al. Prevalence of subclinical coronary artery atherosclerosis in the general population. Circulation. 2021;144(12):916–929. doi:10.1161/CIRCULATIONAHA.121.055340

5. Lindholt JS, Søgaard R, Rasmussen LM, et al. Five-year outcomes of the Danish Cardiovascular Screening (DANCAVAS) trial. N Engl J Med. 2022;387(15):1385–1394. doi:10.1056/NEJMoa2208681

6. von Elm E, Altman DG, Egger M, Pocock SJ, Gøtzsche PC, Vandenbroucke JP; STROBE Initiative. The Strengthening the Reporting of Observational Studies in Epidemiology (STROBE) statement: guidelines for reporting observational studies. Lancet. 2007;370(9596):1453–1457.

7. SydCura. Broward Heart Project Final Report: Report Period May 2023 Through September 2025. Unpublished report prepared for Broward County; January 23, 2026.

8. SydCura. Broward Heart Project: Final Monthly Reporting, Data Collected Through November 30, 2025. Unpublished presentation prepared for Broward County.

9. Cury RC, Leipsic J, Abbara S, et al. CAD-RADS 2.0 – 2022 Coronary Artery Disease-Reporting and Data System: an expert consensus document of the Society of Cardiovascular Computed Tomography (SCCT), the American College of Cardiology (ACC), the American College of Radiology (ACR), and the North America Society of Cardiovascular Imaging (NASCI). J Cardiovasc Comput Tomogr. 2022;16(6):536–557. doi:10.1016/j.jcct.2022.07.002

10. McClelland RL, Chung H, Detrano R, Post W, Kronmal RA. Distribution of coronary artery calcium by race, gender, and age: results from the Multi-Ethnic Study of Atherosclerosis (MESA). Circulation. 2006;113(1):30–37. doi:10.1161/CIRCULATIONAHA.105.580696

11. Detrano R, Guerci AD, Carr JJ, et al. Coronary calcium as a predictor of coronary events in four racial or ethnic groups. N Engl J Med. 2008;358(13):1336–1345. doi:10.1056/NEJMoa072100

12. US Census Bureau. QuickFacts: Broward County, Florida (population estimates, July 1, 2025, V2025). Accessed September 20, 2026. https://www.census.gov/quickfacts/browardcountyflorida

